# The Use of Home-Based Behaviours for Detecting Early Dementia: *Protocol for the CUBOId Study*

**DOI:** 10.1101/2024.02.21.24303130

**Authors:** James Selwood, Niall Twomey, Ian Craddock, Liz Coulthard, Daniel Kumpik, Margaret Newson, Rafael Poyiadzi, Raul Santos-Rodriguez, Weisong Yang, Yoav Ben-Shlomo

## Abstract

**Introduction:** There is a pressing need to automatically understand the state and progression of chronic neurological diseases such as dementia. The emergence of state-of-the-art sensing platforms offers unprecedented opportunities for indirect and automatic evaluation of disease state through the lens of behavioural monitoring. The ContinUous behavioural Biomarkers Of cognitive Impairment (CUBOId) project specifically seeks to characterise behavioural signatures of mild cognitive impairment (MCI) and Alzheimer’s disease (AD) in the early stages of the disease. Bespoke behavioural models will be introduced and deployed on a novel dataset of longitudinal sensor data from persons with MCI and AD to analyse key symptoms of the disease.

**Methods and analysis:** CUBOId is a longitudinal observational study. Participants have diagnoses of MCI or AD, and controls are their live-in partners with no such diagnosis. Multimodal activity data were passively acquired from wearables and in-home fixed sensors over timespans of 2–22 months. Behavioural testing is supported by neuropsychological assessment for deriving ground truths on cognitive status. Machine learning will be used to generate fused multimodal sensor data for optimisation of diagnostic and predictive performance from localisation, activity, and speech together.

**Ethics and dissemination:** CUBOId was approved by an NHS Research Ethics Committee (Wales REC; ref: 18/WA/0158) and is sponsored by the University of Bristol. It is also supported by the National Institute for Health Research (NIHR) Clinical Research Network West of England. Results will be reported at conferences and in peer-reviewed scientific journals.

## 1 Introduction

Neurodegenerative dementia is a growing health problem, with a worldwide prevalence of over 50 million people (Patterson, 2018) that is projected to reach 139 million by 2050 (Organization., 2021), placing ever greater demand on global health and social care systems (Montero-Odasso et al., 2020). Neurodegenerative dementia causes progressive cognitive and behavioural impairments that significantly impact activities of daily living (ADL) (Saari et al., 2020). Alzheimer’s disease (AD), the most common cause (Prince et al., 2014), is now widely accepted as having a long latency period (Bateman et al., 2012; Braak and Del Tredici, 2015; Hampel et al., 2016), but early neurodegeneration can be difficult to distinguish from normal ageing. This is because cognitive decline in people not necessarily destined to develop dementia can also start in middle age (Singh-Manoux et al., 2012). The most sensitive AD markers are expensive and invasive. For example, amyloid positron emission tomography (PET) imaging of people at risk of AD can show changes 15 years before symptom onset (Bateman et al., 2012). Both methods carry a significant false positive and negative rate, and detecting significant changes can take a year or more (Vandenberghe et al., 2013). Routine cognitive testing, while more practical, is far less sensitive to these early changes, with impaired episodic memory manifesting ten years and Mini Mental State Examination (MMSE) five years before symptom onset (Bateman et al., 2012).

Mild cognitive impairment (MCI) is defined by objective evidence of cognitive deficits but preserved daily functioning. Causes of MCI are multiple and include AD (Petersen et al., 1999). In emerging AD, MCI is a prodrome to further cognitive decline, impairment of ADL performance and conversion to dementia. The annual conversion rate varies from 8-15% per year (Petersen, 2009), reflecting the heterogenous nature of MCI. At 5 years follow-up, approximately 50% of patients with MCI have not declined (Rossini et al., 2020). The challenge is to identify which patients are likely to convert and this has typically been done by measuring biomarkers of AD within the MCI population.

A further limitation is that pathological markers do not reflect real-world changes in patient wellbeing and performance (Howieson, 2019). Cognitive tests such as the MMSE (Folstein et al., 1975) and Montreal Cognitive Assessment (MoCA) (Nasreddine et al., 2005) are widely used but are susceptible to confound from practice and patient-related factors, such as effort, fatigue and mood (Lippa, 2018). In-home sensing technologies have the potential to permit longitudinal disease monitoring, providing greater sensitivity than existing biomarkers (Dodge et al., 2012). A recent systematic review of in-home monitoring of dementia showed walking speed and activity around the home to be the most frequently used indicators of MCI (Lussier et al., 2018). This work lends support to the promising potential for home sensor technology to be used in the early detection of dementia.

The Continuous behavioural Biomarkers Of cognitive Impairment (CUBOId) proposal leverages a national investment of £12 million in the EPSRC Sensor Platform for HEalthcare in a Residential Environment (SPHERE) Interdisciplinary Research Collaboration, which aims to greatly advance the state of the art in sensing for health-related behaviours at home. Sponsored by the University of Bristol (UoB), CUBOId will develop novel computational behavioural analysis algorithms that derive dementia-relevant signatures from SPHERE sensor networks. Through unique SPHERE technology, this project addresses the desperate need for accurate, functionally relevant, early dementia biomarkers that are sensitive to meaningful change. The aim is to identify previously unseen behavioural biomarkers of early AD pathology (Poyiadzi et al., 2020) such as sleep disturbances, partner shadowing, wandering and disrupted conversational speech, derived from autonomous, continuous, long-term measurement in patients’ homes. We plan to validate these against neuroimaging (amyloid PET) data and longitudinal neuropsychological progression as measured by repeated cognitive testing. The patient’s live-in companions are also participants, to act as controls and to investigate interactional biomarkers of cognitive decline. It is anticipated that CUBOId will develop, test and de-risk a unique asset for the future community of researchers in the UK Dementias Research Institute, which may be applied to cohorts in the Dementia Platform UK (DPUK) as well as in the international community.

This is a pilot study into the value of smart home sensing technology for detecting behavioural biomarkers in early MCI and AD. CUBOId has two key objectives: 1) To explore SPHERE home sensing of ADL performance as a biomarker of early AD in people with MCI, comparing sensitivity with that of brain imaging and clinical progression over 12-18 months; and 2) to determine to what degree ADL performance variability detected through home-sensing may be explained by potential contextual variables such as the time of day, sleep and daily habits.

## 2 Methods

### 2.1 Patient and Public Involvement (PPI)

SPHERE has run co-design workshops and has trialled its consent processes with members of the public, who averaged a score of 86% on a test designed to show whether they had properly understood the risks (70% was deemed sufficient) and in this mock exercise 92% of the group consented to be part of the experiment. We conducted pilot experiments of behavioural tasks with CUBOId patients, and a PPI advisory team aided with recommendations on participant recruitment and materials, and will be consulted on the strategy to disseminate results to participants, the public and healthcare professionals.

### 2.2 Participants and Eligibility Criteria

A conventional sample size calculation was not performed as this is a feasibility study with no obvious effect estimates. Patients with a diagnosis of MCI or AD were eligible, provided they had no severe physical or psychiatric comorbidity that would interfere with their ability to take part in the study. Their diagnosis was reviewed at their screening appointment. Participants were selected according to the following criteria:

1. Full capacity to give informed consent;
2. At least 50 years old;
3. Competent English speaker;
4. Normal or corrected-to-normal vision;
5. Consent to have the SPHERE system installed in their home.

Participants with a diagnosis of MCI were offered an MRI brain scan and an amyloid PET scan, unless contraindicated. Patients could still take part if they did not have any neuroimaging carried out. Participants were recruited from the cognitive neurology service at North Bristol NHS Trust, the local Dementia Wellbeing Service and the Research Institute for the Care of Older People (RICE). The national Join Dementia Research (JDR) database was also screened to identify potential participants.

Participants were reimbursed for excess electrical usage caused by the SPHERE equipment, and for any costs in the event that sensor installation or removal damaged the property.

### 2.3 Impact of COVID-19 on Data Collection

CUBOId was fortunate in that most data could be collected passively and remotely, although COVID-19 presented many challenges, particularly with respect to cognitive and behavioural testing, and to installation and maintenance of SPHERE infrastructure. We originally intended to visit participants’ homes to perform cognitive testing and to collect behavioural data across 3-4 testing blocks per participant pair, held at roughly 0, 6 and 12 months after SPHERE installation, with the possibility of a fourth block at 18 months if possible. However, we suspended SPHERE deployments in March 2020 and introduced remote testing protocols to mitigate risk to participants, including extensive revisions of the deployment procedure to minimise the time spent in participants’ homes by CUBOId technicians. The neuropsychological test battery was also adapted for remote data collection during the pandemic. Cognitive assessments were conducted online using the online video-conferencing platform Attend Anywhere^1^. Some tests (the flanker test and the supermarket task) were dropped as they could not be administered online. For CUBOId’s longitudinal speech task (the TV task (Kumpik et al., 2022)), we sent tablet devices to participants by courier and provided training and support with the tasks online or by telephone. All devices were cleaned with disinfectant wipes and participants were instructed to leave them in their packaging for 3-4 days before removing to reduce the risk of viral transmission.

The CUBOId team designed a series of behavioural tasks to test the performance of individuals undertaking various ADL around the home, also planned to take place at baseline, 6 months and 12 months. SPHERE would collect data while the tasks were being performed, allowing comparisons to be made between the SPHERE data and the ground truth obtained from the recording. The first task required the participant to make a cup of tea in a specific manner with verbal instructions of varying complexity. The second task required the participant to complete a series of simple everyday tasks in different rooms around the home, such as filling the kettle with water or drawing the curtains. The third task was a repetition of this sequence of behaviours under high cognitive load, i.e., while performing a phonemic fluency test. The tasks were intended to be administered by a researcher visiting the participant’s home, but were suspended due to the pandemic.

### 2.4 Neuropsychological and Cognitive Tasks

Participants completed neuropsychology testing throughout the study to determine whether the data collected by SPHERE is more sensitive to change than cognitive tests used in dementia research trials. The test battery was adapted from the recommendations of the European Prevention of Alzheimer’s Dementia (EPAD) study (Ritchie et al., 2017). The tests (see Table 1) are administered by a clinical fellow.

**Table 1:** Test battery recommended in the EPAD study. Table adapted from Ritchie et al. (2017).

| Cognitive Domain | Test | Platform |
| --- | --- | --- |
| Reaction time/information processing speed/conceptual shifting/selective attention | Coding | RBANS |
| Verbal episodic memory | List learning | RBANS |
|  | Story memory | RBANS |
| Visuospatial analysis | Figure copy | RBANS |
|  | Line orientation | RBANS |
| Language | Picture naming | RBANS |
|  | Semantic fluency | RBANS |
| Working memory | Digital span | RBANS |
|  | Dot counting | NIH examiner/toolbox |
| Allocentric space | Four mountains task | iPad app |

In addition to these tests, participants also completed the Alzheimer’s Disease Assessment Scale – Cognitive subscale (ADAS-Cog (Kueper et al., 2018)). The aim was to determine whether the EPAD tests and/or SPHERE data analyses are more sensitive to change than the ADAS-Cog. The Test Of Memory Malingering (TOMM) is a 50-item visual recognition test used at baseline as a test of performance validity (Tombaugh, 1996).

Participants also completed a series of questionnaires to assess their psychological wellbeing and sleep. The 6-Item State-Trait Anxiety Inventory (Marteau and Bekker, 1992) and Geriatric Depression Scale (GDS; short version (Sheikh and Yesavage, 2014)) assess for anxiety and depression. Participants also completed the Epworth Sleepiness Scale (Johns, 1991) and the Pittsburgh Sleep Quality Index (Buysse et al., 1989). If the GDS or ESS suggest clinically significant depression or excessive daytime sleepiness, then this was assessed further and the general practitioner (GP) was informed, if appropriate.

Some participants have hearing problems, a known risk factor for cognitive decline. We will therefore issue participants with a custom hearing questionnaire derived from the Speech, Spatial and Qualities of Hearing questionnaire (short form) (Moulin et al., 2019) and the Glasgow Hearing Aid Benefit Profile (Gatehouse, 2000) (see supplemental material S1 in (Kumpik et al., 2022) for details).

Finally, participants are invited to complete a gamified smartphone memory task at home (Mezurio (Lancaster et al., 2020)). Participants completed the task daily for one month at baseline, 6 months and 12 months follow-up.

### 2.5 Neuroimaging

All patients with MCI were invited to have an MRI brain scan and an amyloid PET scan to discriminate incipient AD from other causes of MCI, such as vascular disease. MRI was used to investigate the macroscopic structure of the brain with post-processing to determine whole brain and hippocampal volumes. The amyloid PET scan findings were only disclosed to patients if they wished to know their amyloid status after appropriate counselling.

### 2.6 Acquiring Natural Conversations - The TV Task

It is well established that language function can be affected early in dementia (Eyigoz et al., 2020; Iacono et al., 2009; Snowdon, 1997; Snowdon et al., 1996). A naturalistic speech task (Kumpik et al., 2022) has been designed to record audio of spontaneous conversations in the home environment using a tablet device. Participants and their study partners were asked to watch a television programme and talk about it in the style of the popular Channel 4 television series, Gogglebox. The conversations will be subjected to linguistic analysis using machine learning, with the aim of detecting participants with and without cognitive impairment.

### 2.7 Multimodal Sensing with SPHERE

The SPHERE collaboration has developed a novel low-power sensing platform for continuous monitoring of information about the home (e.g., temperature, humidity and energy use), and of information about the health-related behaviours of people within the home (e.g., location, activity levels, locomotion and sleep patterns). The technologies were developed in a prototype smart house (Figure 1), which has been fitted with a range of environmental, video and wearable sensors (Woznowski et al., 2017). The following subsections describe the sensing modalities that were deployed in the smart home. All sensors are synchronised with Network Time Protocol.

**Fig. 1:**
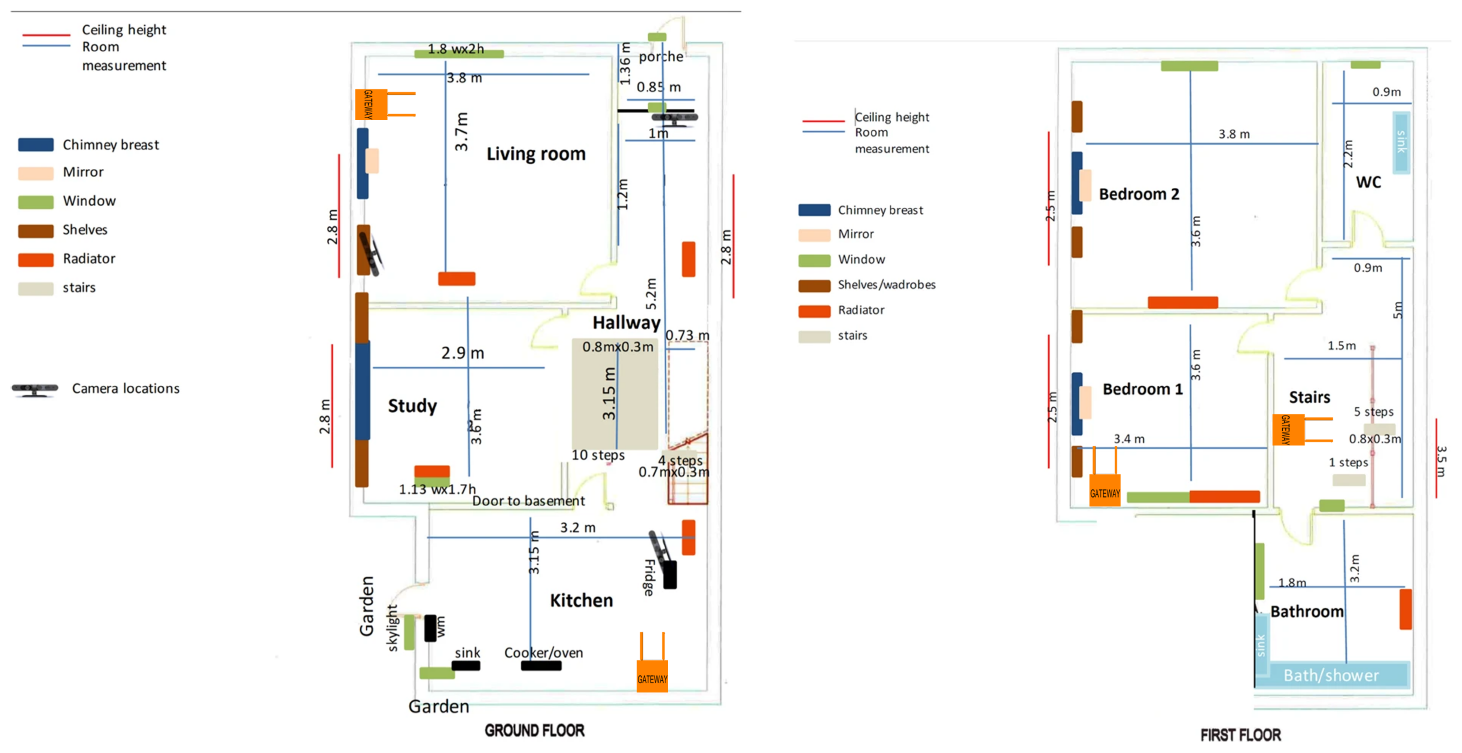
Floor plan of the prototype SPHERE house.

#### Wearable Accelerometers

Participants wore the SPHERE wearable (Figure 2) on their dominant wrist. The wearable was an acceleration-based activity sensor, equipped with two ADXL362 accelerometers, which wirelessly transmitted data using the BLE standard to several access points positioned within the house (Fafoutis et al., 2017). The sensor outputs are continuous numerical accelerometer readings (units of g, i.e., approximately 9.81 *m/s*^2^).

**Fig. 2:**
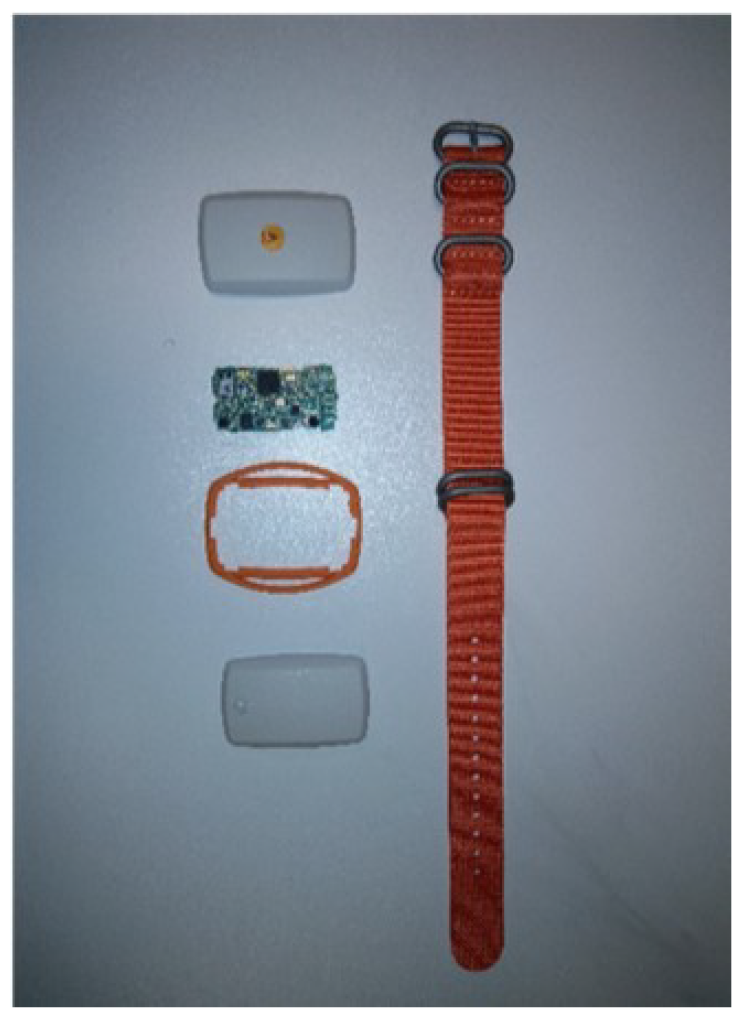
The SPHERE wearable.

The wearables also provided received signal strength Indicators (RSSI) recorded at each access point (in dBm) for indoor localisation of participants. The accelerometers recorded data at 27 Hz (12-bit resolution), and the accelerometer ranges were between *±* 4*g*. RSSI values are also recorded at 5 Hz, and values are no lower than -128 dBm.

Designed for long-term operation with minimal maintenance, the system was optimised for low energy consumption. To that end, the wearable sensor communicated with the smart house using undirected connectionless BLE advertisements. Although data reliability can be addressed at the receiver (Fafoutis et al., 2017), this communication approach does not guarantee data packet delivery. Nevertheless, recent work by SPHERE researchers demonstrates robust activity recognition using accelerometer data (Diethe et al., 2015; Twomey et al., 2018). Data from the SPHERE wearable have also been used for the validation of a privacy-preserving algorithm for wearable embedded systems (Fafoutis et al., 2016).

#### Silhouette Sensors

SPHERE incorporates 3D ‘silhouette cameras’ in its sensor network. These work by broadcasting patterns of infrared light over the camera’s field of view and measuring the light’s reflection latency. This allows a point cloud of depth to be derived and added to the video information.

One camera was installed in the kitchen. To preserve privacy and anonymity only the coordinates of the 2D and 3D bounding boxes and silhouette centres of mass were stored (Figure 3). Bounding boxes are the minimal axis-aligned 2D rectangles and 3D cubes that enclose the silhouette and the depth point cloud of the detected participants. No raw video data was stored.

**Fig. 3:**
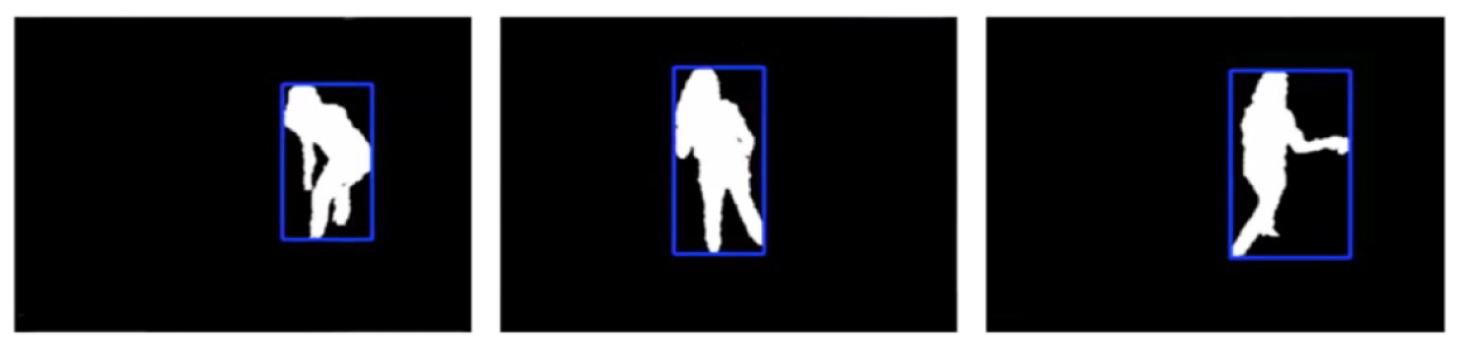
Silhouette sensing in SPHERE: Images from 3D infrared cameras are reduced to non-identifiable silhouettes and bounding boxes. Here a person is shown cooking.

We used ASUS Xtion PRO RGB-D cameras^2^, with automatic detection of participants achieved using the OpenNI library^3^. Post-processing methods were employed to minimise false positives.

#### Environmental Sensors

Whereas the wearable and silhouette sensors measured features relating to individuals, the environmental sensors captured features relating to wider ambient aspects of the home environment. The environmental sensors deployed in SPHERE are:

- **Passive infrared (PIR):** These measure infrared light radiating from objects in rooms to detect the presence of humans.
- **Temperature:** For measuring ambient room temperature in °C.
- **Humidity:** For measuring room humidity as a percentage between 0% and 100% air moisture saturation.
- **Barometric pressure:** Measures the barometric pressure of a room in hectopascals (hPa).
- **Light level:** Reports a room’s current light level in lux.
- **Power usage:** Records the electricity usage of appliances from their wall sockets. Appliances are typically equipped with power sensors including the kettle, microwave, oven, toaster and television.
- **Water flow:** This reports the status of the cold and hot taps (‘on’/’off’).

With the exception of the water flow and power sensors, the environmental sensors were housed in one module (Figure 4), with one module deployed per room. The light sensor was also enclosed within the housing unit, but still yielded meaningful changes in light.

**Fig. 4:**
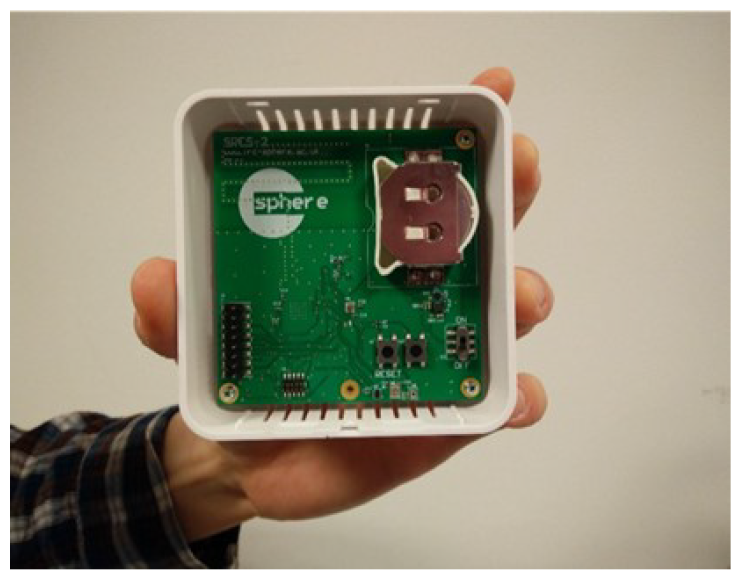
Housing unit containing the environmental sensor package.

### 2.8 Platform Installation

Sensors were installed by trained technicians. At the time of writing, we have installed SPHERE sensor networks in over 50 homes in the Bristol, UK region. There were four main visit types by the installation team to a home:

1. **Home evaluation** This served as an introduction to the SPHERE team that would manage the sensor installation. The floor plan of the home was recorded (including locations of key appliances, sockets, and high-use rooms), and the deployment team then decided on the sensor deployment locations.
2. **Sensor installation** During this visit, the sensors were installed. Typically, one PIR sensor was installed in each main room of the house. One silhouette sensor was installed in the kitchen. Electricity monitors were installed on key appliances (e.g. microwave, television), and up to two water meters were installed (one in the kitchen sink and one in the main bathroom). Some flexibility to this typical installation was required depending on individual house characteristics.
3. **Maintenance visits** In the event of sensor problems the technical team visited the home, diagnosed the problem and implemented repairs. Maintenance visits were sometimes also needed for critical security updates, although these could sometimes be applied remotely.
4. **SPHERE removal** This was the final visit to a home for the removal of the sensor system.

### 2.9 Data Analysis

The data analysis strategy for CUBOId’s naturalistic conversational speech task (TV task) has been detailed elsewhere (Kumpik et al., 2022).

To validate smart home-derived biomarkers of cognitive decline, data analysis will examine pre-defined research questions specific to relevant behaviours. This will be complemented by exploratory data analyses to identify novel relevant behavioural traits. We will use machine learning to analyse both behaviours that occur regularly and are affected by cognitive decline (such as increased sedentary behaviour), and those targeting specific symptoms of AD (e.g. wandering, partner shadowing). Longitudinal analyses will identify both outlying instances (i.e., short periods of unusual behaviour) and longer-term changes. One of our core hypotheses is that the impact of cognitive decline in a household will affect the behaviour of both the patient and their study partner. These analyses will therefore also be performed on data from the study partner. In the following sections, we describe pre-defined behaviours we have identified as likely to be sensitive to cognitive decline.

#### Behaviours Relevant to Cognitive Decline

- **Room occupancy:** Since some ADL take place exclusively in a particular room (e.g., cooking in the kitchen) the distribution over rooms occupied by persons with progressive cognitive decline may change over time. The wearable’s RSSI readings and the PIR sensor data will be used to quantify room occupancy distributions throughout data collection.
- **Wandering and partner shadowing:** The wearables and PIR sensors will help detect two behaviours characteristic of AD: random and purposeless wandering from room to room, and shadowing (e.g., following a partner around the house to seek comfort).
- **Sleep disturbance:** People with MCI or dementia usually suffer from different levels of sleep disturbance. The wearables and PIR sensors installed in the bedrooms of the CUBOId’ houses will also be used to study the sleep patterns of participants.

## 3 Ethics and Dissermination

While all participants were required to have the capacity to consent at the beginning of the study, the possibility of losing capacity during the study could not be excluded. On occasions where this happened the study partner was approached to decide whether the participant should continue or not according to previously stated preferences. If it was agreed that the participant would wish to continue, he/she was only expected to continue with the SPHERE monitoring and did not need to complete any other components of the CUBOId protocol.

CUBOId is sponsored by UoB and was approved by the Wales Research Ethics Committee (REC) 7 (ref: 18/WA/0158) and the Health Research Authority (IRAS project ID 234027). CUBOId is supported by the National Institute for Health Research (NIHR) Clinical Research Network West of England. Protocol amendments and adverse events are referred to the REC and UoB. UoB has Public Liability and Professional Negligence insurance policies to cover the eventuality of harm to a research participant or University employee arising from the design or management of the research. Unforeseen protocol deviations are reported to the Principal Investigator, and serious deviations to the Sponsor, immediately. If CUBOId is ended prematurely, the Principal Investigator will notify the UoB and Wales RECs, including the reasons for termination.

## 4 Data Statement

All individuals in the CUBOId team have unrestricted access to the full dataset. Results will be reported at conferences, in peer-reviewed scientific journals and on the UoB website. Both the anonymised data and study documentation are stored on secure servers within UoB and shared according to UoB procedures and guidelines. Personally identifiable information is stored separately to research data. Data management procedures follow the stipulations of the General Data Protection Regulation. In line with NIHR guidance, we obtained consent for participants’ data to be stored for up to 20 years and shared anonymously with other researchers, after appropriate review to ensure it will be used according to the ethics approval.

## Data Availability

All data produced in the present study are available upon reasonable request to the authors.

## Acknowledgements

We would like to thank Julia Carey, David Bailey and Danielle Hale for their support in administering CUBOId and in collecting data. We would also like to thank our participants for their involvement in CUBOId.

## Footnotes

1 https://www.attendanywhere.com

2 https://www.asus.com/3D-Sensor/XtionPRO/

3 https://github.com/OpenNI/OpenNI

